# Cost-Effectiveness of Integrative Concepts for the Treatment of Oncologcial Patients (OCEANic): Protocol for a Systematic Review

**DOI:** 10.1101/2025.05.02.25326858

**Authors:** Friedemann Schad, Anja Thronicke, Thomas Reinhold, Shiao Li Oei, Hiba Boujnah, Elisa Baldini, Henrik Szóke, Nina Fuller-Shavel, Georg Seifert

## Abstract

**Background:** Integrative oncology combines conventional cancer treatments with complementary medicine to enhance patient-centered care, symptom management, and overall well-being. Despite growing demand in Europe, challenges such as unequal access, reimbursement policies, and limited cost-effectiveness research hinder its broader integration into healthcare systems. This systematic review will evaluate the financial sustainability of integrative oncology interventions, providing evidence to support policy decisions and future investments in patient-centered cancer care.

**Methods:** A systematic review will be conducted following PRISMA guidelines, applying PICO criteria for study inclusion, focusing on adult cancer patients receiving integrative therapies and evaluating their economic impact. The review is currently in the literature search phase, using predefined search terms across selected databases. Studies published in peer-reviewed journals, including randomized and non-randomized trials, will be considered, while studies on adolescents or without economic evaluations will be excluded. Records will be managed using EndNote, and two independent reviewers will assess study eligibility through title and abstract screening, resolving discrepancies through discussion, with a third reviewer if needed. Data will be extracted independently using a standardized form, including study characteristics, sample size, outcomes, and cost-effectiveness details. Comparative analyses will focus on clinical and humanistic outcomes, using key metrics like ICER and CBR. A narrative synthesis will evaluate study quality and methodology, with no meta-analysis due to expected heterogeneity. As this study does not involve human participants or personal data, ethics approval is not required.

**Results:** The systematic review is currently in progress. Pilot work and initial study identification have begun, though not yet completed. Screening of search results is scheduled to commence in mid-May 2025, followed by data extraction in early June. Completion of the review is anticipated by the end of October 2025.

**Discussion:** The findings will provide valuable insights into the cost-effectiveness of integrative oncology interventions, informing healthcare policy, clinical practice, and future research directions. By identifying financially sustainable integrative treatments, this project aims to enhance patient care and optimize resource allocation within oncology healthcare systems.

**Trial registration:** PROSPERO, registration number 1019386, date of registration 25 March, 2025.

## Introduction

### Rationale [6]

Integrative oncology (IO) is defined as “*Integrative oncology is a patient-centered, evidence-informed field of cancer care that utilizes mind and body practices, natural products, and/or lifestyle modifications from different traditions alongside conventional cancer treatments*” (3). IO is gaining recognition for improving patient-centered care, addressing quality of life, symptom management, and overall well-being. In Europe, there is increasing interest and demand from citizens for integrative oncology services, as well as successful examples of meaningful integration of conventional medicine with Traditional, Complementary, and Integrative Healthcare (TCIH). However, access to TCIH practitioners, the availability of medicinal products, and reimbursement policies remain uneven. These challenges hinder the fulfillment of patients’ rights to choose their preferred healthcare approaches. Cost-effectiveness in this field remains underexplored, making it challenging to determine their financial sustainability and feasibility within healthcare systems. Assessing the cost-effectiveness of integrative oncology is particularly relevant given the financial constraints faced by healthcare systems across Europe.

Systematic reviews in the field of integrative oncology are rare. An economic evaluation by Huebner et al. emphasized the challenges in conducting economic evaluations of complementary and alternative medicine in oncology (5). The authors pointed out difficulties in defining effectiveness parameters and selecting appropriate comparative treatments. They recommended a comprehensive assessment approach that incorporates both direct and indirect costs, tailored to the holistic nature of complementary and alternative therapies. In summary, while existing literature indicates that integrative oncology may offer cost-effective benefits, the cumulative evidence is limited and varies in quality. Further rigorous economic evaluations are necessary to draw definitive conclusions about the cost-effectiveness of integrative approaches in cancer care. Most systematic reviews on the cost-effectiveness of integrative medicine tend to focus on broader populations rather than specifically on oncological patients. For instance, a comprehensive systematic review by Herman et al. analyzed 338 economic evaluations of complementary and integrative medicine up to 2010 (4). Among the 114 full economic evaluations published between 2001 and 2010, 27% met high-quality criteria, and 29% of these demonstrated health improvements alongside cost savings compared to usual care. However, the authors noted significant variation in the overall quality of these studies and recommended more rigorous research to support these findings. Additionally, a systematic review currently registered in 2024 in the PROSPERO registry is still ongoing and has yet to be finalized. While it encompasses all types of complementary, alternative, traditional, and conventional medicine, it does not focus on oncological patients and specifically evaluates the economics of these therapies in low- and middle-income countries (11).

### Objective [7]

This study protocol describes a systematic review that assesses cost-effectiveness of integrative oncology care. The systematic review will identify key cost drivers, evaluate methodologies used in cost-effectiveness analyses, and explore the feasibility of integrating complementary and alternative treatments into conventional oncology care. The findings will support evidence-based decision-making for policymakers, healthcare providers, and researchers, guiding future investments in patient-centered cancer care and promoting the adoption of integrative oncology at European, national, and regional levels.

## METHODS

A systematic review will be conducted in accordance with the PRISMA guidelines to ensure a rigorous and transparent methodology (2)]. Eligibility criteria according to PICO framework (Population, Intervention, Comparison, Outcome) will be applied (6). A 17-item checklist in accordance with Moher et al. has been included as supplementary table S1.

### Eligibility criteria [8]

#### Inclusion criteria

Only studies published in peer-reviewed journals will be considered. The systematic review will include randomized controlled as well as non-randomized studies. **Population**: Single studies involving data of adults diagnosed with oncological primary disease will be included. **Intervention**: Eligible studies must involve data on cancer patients receiving integrative therapies. Studies in which a comparison between at least two groups has been performed and one of these groups has received integrative oncological treatment (search terms for integrative oncology can be found in chapter “Search strategy”). Pre-clinical studies will be excluded. **Comparison:** Studies will be included in which at least one integrative oncology concept is compared to at least one conventional, standard-oncological or guideline-oriented oncological concept (which may include watch-and wait strategies as well). **Outcome:** This review will include studies that evaluate the cost-effectiveness, cost-benefit, cost-utility, or economic impact of integrative oncology interventions. **Perspective:** The systematic review must use at least one recognized perspective—for example, society, third-party payer, hospital or employer.

#### Exclusion criteria

Studies that do not include an economic evaluation component, as well as abstracts, conference proceedings, and non-peer-reviewed sources, will be excluded. Additionally, studies that focus on integrative medicine but are not related to cancer care will not be considered for inclusion.

### Information Sources [9]

The search will employ a comprehensive and sensitive topic-based strategies designed for each database from inception to 7 April, 2025. There will be no language or geographical restrictions. Databases include CINAHL – Cumulative Index to Nursing and Allied Health Literature, EconLit, and EMBASE vis Ovid, PsycInfo, and PubMed.

### Search strategy [10]

The search will incorporate Boolean search strings using key terms related to integrative oncology, health economic evaluation, and oncology treatment models, see search strategy, table 1. Other studies will be identified by contacting authors or experts, looking through all the articles that cite the papers included in reviews and searching trial or study registers. Only published studies will be sought. There are no language and no data restrictions. The search will be performed by two independent researchers.

**Table 1.**
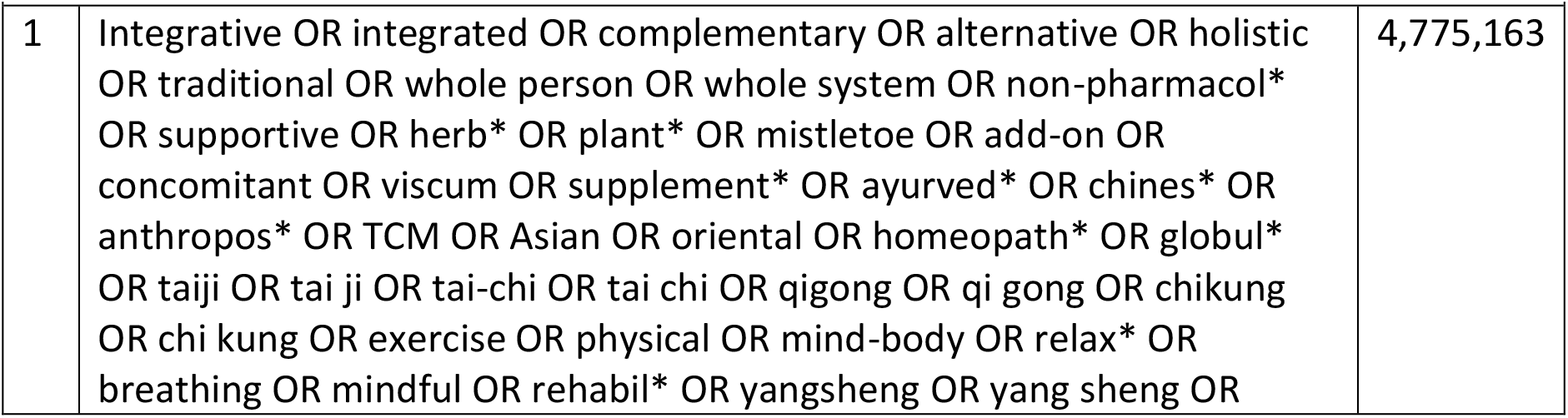
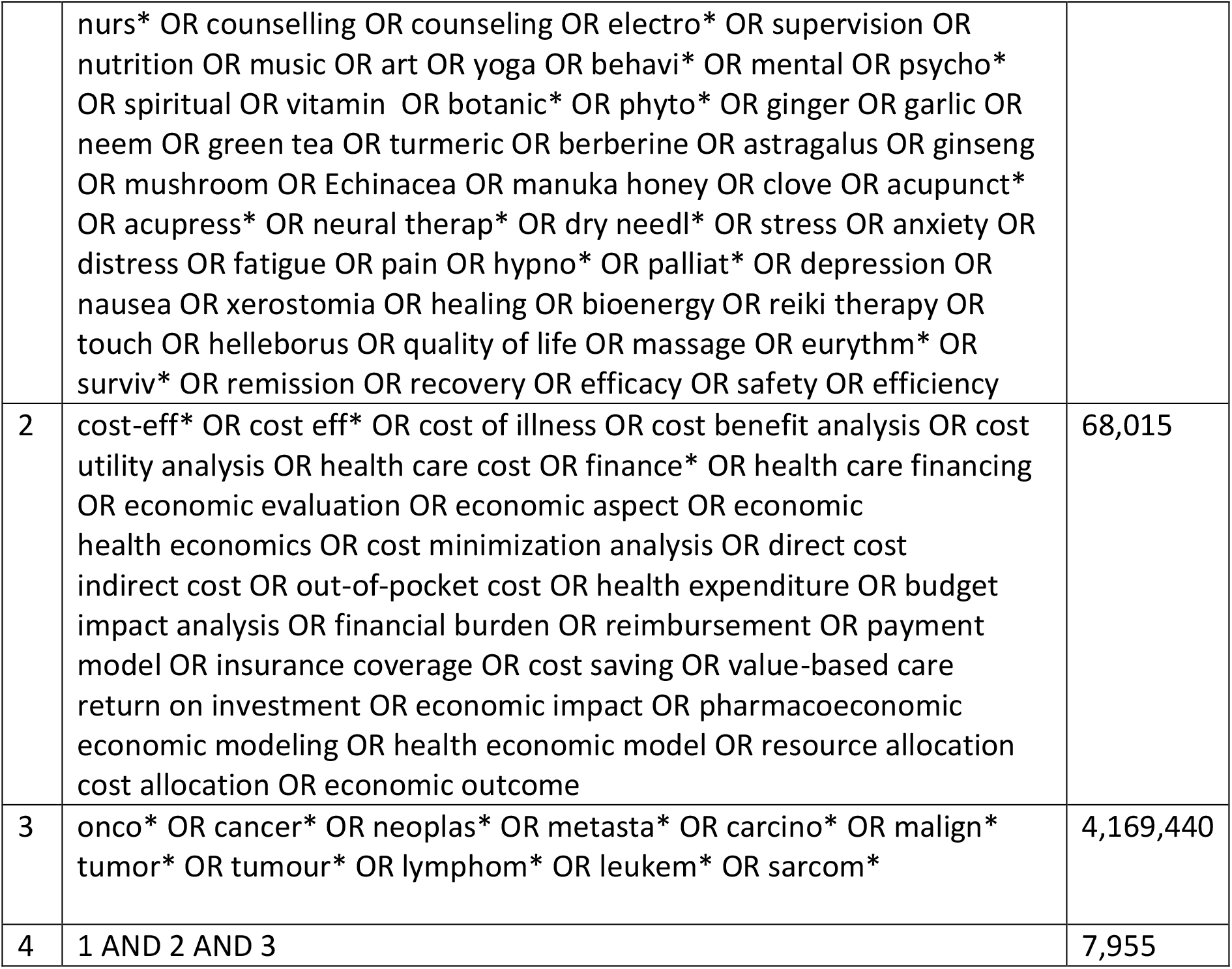
Boolean search strategy; advanced search 08.04.2025.

| Table 1.<br>Boolean search strategy; advanced search 08.04.2025 |  |  |
| --- | --- | --- |
| 1 | Integrative OR integrated OR complementary OR alternative OR holistic OR traditional OR whole person OR whole system OR non-pharmacol* OR supportive OR herb* OR plant* OR mistletoe OR add-on OR concomitant OR viscum OR supplement* OR ayurved* OR chines* OR anthropos* OR TCM OR Asian OR oriental OR homeopath* OR globul* OR taiji OR tai ji OR tai-chi OR tai chi OR qigong OR qi gong OR chikung OR chi kung OR exercise OR physical OR mind-body OR relax* OR breathing OR mindful OR rehabil* OR yangsheng OR yang sheng OR | 4,775,163 |
|  | nurs* OR counselling OR counseling OR electro* OR supervision OR nutrition OR music OR art OR yoga OR behavi* OR mental OR psycho* OR spiritual OR vitamin OR botanic* OR phyto* OR ginger OR garlic OR neem OR green tea OR turmeric OR berberine OR astragalus OR ginseng OR mushroom OR Echinacea OR manuka honey OR clove OR acupunct* OR acupress* OR neural therap* OR dry needl* OR stress OR anxiety OR distress OR fatigue OR pain OR hypno* OR palliat* OR depression OR nausea OR xerostomia OR healing OR bioenergy OR reiki therapy OR touch OR helleborus OR quality of life OR massage OR eurythm* OR surviv* OR remission OR recovery OR efficacy OR safety OR efficiency |  |
| 2 | cost-eff* OR cost eff* OR cost of illness OR cost benefit analysis OR cost utility analysis OR health care cost OR finance* OR health care financing OR economic evaluation OR economic aspect OR economic health economics OR cost minimization analysis OR direct cost indirect cost OR out-of-pocket cost OR health expenditure OR budget impact analysis OR financial burden OR reimbursement OR payment model OR insurance coverage OR cost saving OR value-based care return on investment OR economic impact OR pharmacoeconomic economic modeling OR health economic model OR resource allocation cost allocation OR economic outcome | 68,015 |
| 3 | onco* OR cancer* OR neoplas* OR metasta* OR carcino* OR malign* tumor* OR tumour* OR lymphom* OR leukem* OR sarcom* | 4,169,440 |
| 4 | 1 AND 2 AND 3 | 7,955 |

## Study records

### Data management [11a]

Records will be managed through End Note, a specific software for managing bibliographies.

### Selection process [11b]

Two independent reviewers (AT and SLO) will independently search information sources and screen titles and abstracts to assess study eligibility. Each eligibility criterion will be graded as *eligible, not eligible*, or *might be eligible* (7). Studies will be considered potentially relevant when they cannot be clearly excluded based on their title and abstract alone (8), following discussion between the two reviewers. Full texts will be obtained for abstracts lacking sufficient information or where there is disagreement. Both reviewers will then independently assess the full text to determine final inclusion based on predefined criteria. Exclusion reasons will be recorded in a standardized data extraction excel-based form. The reviewers will compare their assessments, resolving discrepancies through discussion. In the event of disagreement, a third reviewer (FS) will assist in the final decision (9).

### Data collection process [11c]

Data will be extracted independently by at least two reviewers (AT/SLO) with a process to resolve differences (8). Authors will be asked to provide any required data not available in published reports. Data will be checked for consistency and clarity. The standardized form will be used to collect information on study characteristics, including study design, year of publication, location, sample size, cancer type, integrative oncology care type, economic-effectiveness evaluation type and cost perspective.

### Data items [12]

Data extracted will include the following summary data: study design, sample characteristics, sample size, outcomes, and timescales to reflect disorder state, acute, subacute and chronic, outcomes.

### Outcomes and prioritization [13]

Comparative analyses will evaluate both costs and outcomes, including clinical and humanistic impacts. Key measures such as the incremental cost-effectiveness ratio, cost-benefit-ratio, and other metrics that compare costs with clinical outcomes, will be included to assess the value of interventions.

### Risk of bias in individual studies [14]

Risk of bias for each included trial will be independently assessed by two independent reviewers (AT/SLO). The quality of the studies will be reviewed using the International Society for Pharmacoeconomics and Outcome Research (ISPOR) Consolidated Health Economic Evaluation Reporting Standards (CHEERS) (10). Data will be assessed with a process to resolve differences. Risk of bias due to missing results will be assessed. Additional information will be sought from study investigators if required information is unclear or unavailable in the study publications/reports. Certainty of findings will not be assessed.

## Data

### Synthesis [15]

Due to the expected heterogeneity of economic evaluations, a meta-analysis will not be conducted. Instead, a rigorous narrative synthesis will evaluate study quality, methodological differences, and robustness. Studies will be systematically described and grouped based on PICO criteria, see chapter “Eligibility criteria”. Any discrepancies in interpretation will be resolved through discussion between reviewers. Studies will also be categorized according to methodology and strategy type. Key data will be tabulated for clarity, and relationships between studies will be analyzed qualitatively, focusing on methodological variations, sensitivity analyses, and potential biases. According to the given data availability patient’s subgroups by diagnosis, age, gender and by type of treatment will be separately presented additionally.

### Confidence in cumulative evidence [17]

For trial-based economic evaluations the Grading of Recommendations, Assessment, Development and Evaluation (GRADE) – approach including risk of bias assessment will be used. Findings on incremental costs/effects, cost-effectiveness ratios and uncertainty measures will be added to the evidence profiles in the respective grade-tables. In addition, results will be plotted into a cost-effectiveness plane in case common metrics (e.g. costs and quality – adjusted life years - QALYs) for included studies are applied.

## Supporting information

PRISMA-P 2015 Checklist, adapted for the use with protocol submissions to Systematic Reviews from Moher et al. 2015, Table 3 (12)

## Contributions [3b]

EB, HB, HS, FS, and GS provided funding searches. AT drafted the manuscript. TR contributed to the health economic issues. All authors contributed to the development of the selection criteria, the risk of bias assessment strategy and data extraction criteria. AT, SLO developed the search strategy and provided statistical expertise. All authors reviewed the search strategy and gave approval. All authors read, provided feedback and approved the final manuscript.

## Amendment [4b]

The current protocol version is version 1.0, dated from 25 March 2025. Important protocol amendments post registration will be recorded and included in dissemination.

## Sponsor/Funder [5b]

This study is being conducted by the EUROCAM, the Charité Competence Center for Traditional and Integrative Medicine (CCCTIM) Charité – Universitätsmedizin Berlin, the Research Institute Havelhöhe at the hospital Havelhöhe (FIH), the IVAA, and the Institute for Social Medicine, Epidemiology and Health Economics Charité. The IVAA, the CCCTIM and the FIH are the sponsors of the systematic review.

The International Federation of Anthroposophic Medical Associations (IVAA), the Weleda AG, the Iscador AG, the Abnoba GmbH, the Helixor Heilmittel GmbH, the Holistic Medicine Network in Berlin, the Lukas GmbH, Labo-Life, and Alpen Pharma GmbH funded this research.

## Role of sponsor/funder [5c]

The sponsor is non-commercial. The sponsor ensures quality management, qualified and trained personnel, protocol compliance, and timely procurement of the systematic review, supports the systematic review with personnel resources, and is involved in the development of the systematic review strategy, methodology and project development. The funders has no role in the development of the protocol or of the systematic review.

## Additional information

### Review Status

The review is currently planned and ongoing. Pilot work as well as formal searching as well as study identification have started but not completed. Screening search results against inclusion criteria is planned to start mid of May 2025, data extraction in the beginning of June 2025, evaluation of risk of bias, quality assessment, and data synthesis are planned to start in July 2025. Writing of the findings is planned in August 2025 and dissemination to a peer-reviewed journal in September. The systematic review is planned to be finished at the end of October 2025. The current protocol version is 1.0, dated 25 March 2025.

### Dissemination Plan

The findings (and the study protocol) of this systematic review will be disseminated through submission to a peer-reviewed journal, presentations at relevant academic and professional conferences, and engagement with policymakers and healthcare providers. The goal is to facilitate informed decision-making on the cost-effectiveness of integrative oncology interventions and to support their integration into cancer care policies.

## List of abbreviations

PROSPERO: International Prospective Register of Systematic Reviews
PRISMA-P: PRISMA for Systematic Review Protocol
CCCTIM: Charité Competence Center for Traditional and Integrative Medicine
FIH: Research Institute Havelhöhe
IVAA: International Federation of Anthroposophic Medical Assocations
IO: integrative oncology
TCIH: Traditional, Complementary, and Integrative Healthcare
PICO: Population, Intervention, Comparison, Outcome
CINAHL: Cumulative Index to Nursing and Allied Health Literature
ISPOR: International Society for Pharmacoeconomics and Outcome Research
CHEERS: ISPOR Consolidated Health Economic Evaluation Reporting Standards
ICER: cost-effectiveness ratio
CBR: cost-benefit ratio.

## Declarations

### Authors approval

All authors have seen and approved the manuscript.

### Patient and public involvement

Patients and/or the public were not involved in the design, or conduct, or reporting, or dissemination plans of this research.

### Data Availability Statement

Any material required to support the protocol can be supplied on reasonable request.

## Acknowledgement

We would like to acknowledge Tido von Schoen - Angerer, Adrianne Waldt, and Almut Altmann involved in the OCEANic project in supporting the present work.

## Supplementary figures and tables

**Supplementary table S1.**
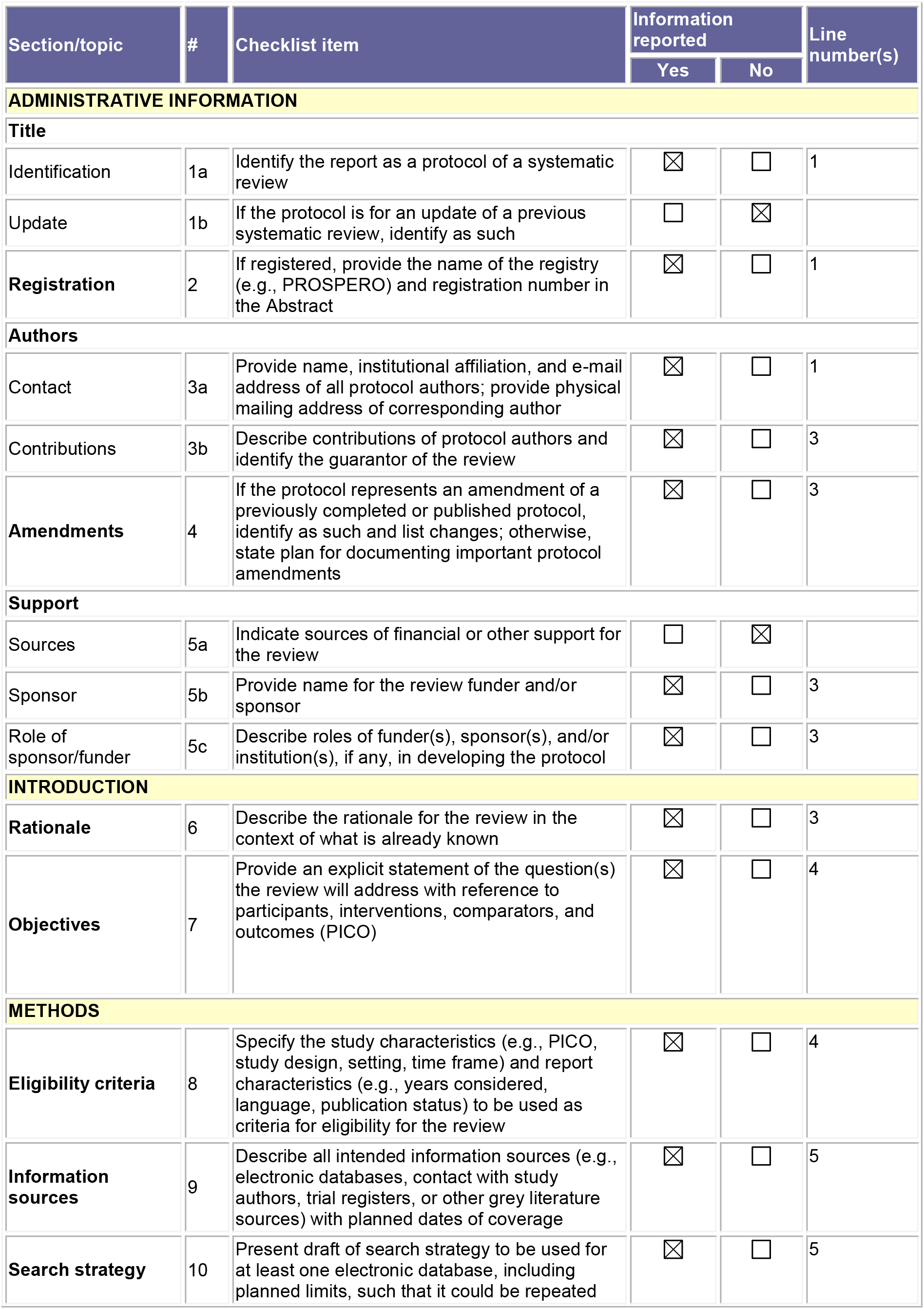
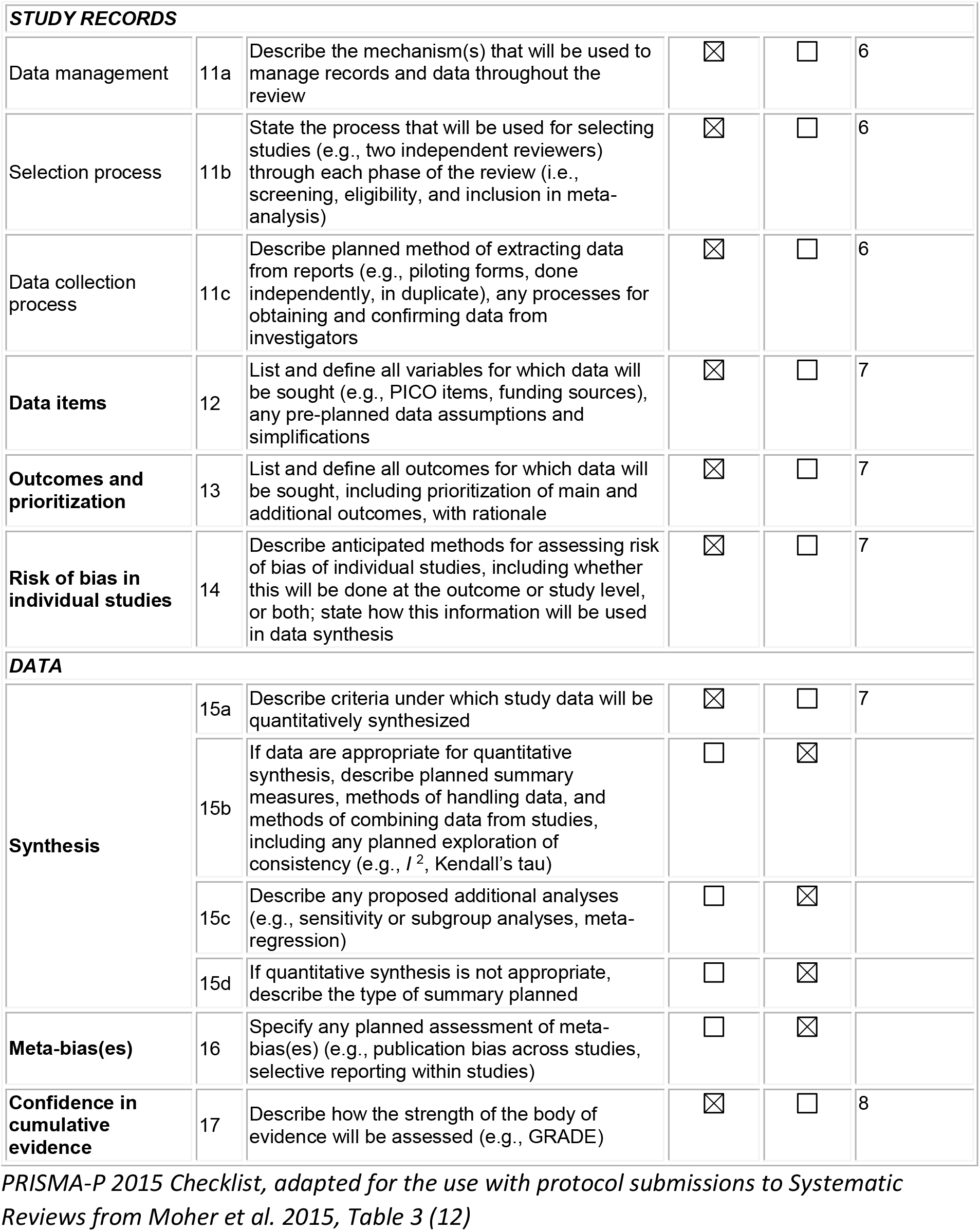
PRISMA-P 2015 Checklist.

## References

1) Schad F, Thronicke A, Oei SL, Reinhold T, Boujnah H, Baldini E, Szoke H, Waldt A, von Schoen-Angerer T, Seifert G. Cost-Effectiveness of Integrative Concepts for the Treatment of Oncological Patients (OCEANic): a Systematic Review. International prospective register of systematic reviews – PROSPERO. Registration ID 1019386. https://www.crd.york.ac.uk/PROSPERO/

2) Shamseer L, Moher D, Clarke M, Ghersi D, Liberati A, Petticrew M, Shekelle P, Stewart LA; PRISMA-P Group. Preferred reporting items for systematic review and meta-analysis protocols (PRISMA-P) 2015: elaboration and explanation. BMJ. 2015 Jan 2;350:g7647. doi: 10.1136/bmj.g7647. Erratum in: BMJ. 2016 Jul 21;354:i4086. doi: 10.1136/bmj.i4086. PMID: 25555855.

3) Witt CM, Balneaves LG, Cardoso MJ, Cohen L, Greenlee H, Johnstone P, Kücük Ö, Mailman J, Mao JJ. A Comprehensive Definition for Integrative Oncology. J Natl Cancer Inst Monogr. 2017 Nov 1;2017(52). doi: 10.1093/jncimonographs/lgx012. PMID: 29140493.

4) Herman PM, Poindexter BL, Witt CM, Eisenberg DM. Are complementary therapies and integrative care cost-effective? A systematic review of economic evaluations. BMJ Open. 2012 Sep 3;2(5):e001046. doi: 10.1136/bmjopen-2012-001046. PMID: 22945962; PMCID: PMC3437424.

5) Huebner J, Prott FJ, Muecke R, Stoll C, Buentzel J, Muenstedt K, Micke O; Prevention and Integrative Oncology of the German Cancer Society Working Group. Economic Evaluation of Complementary and Alternative Medicine in Oncology: Is There a Difference Compared to Conventional Medicine? Med Princ Pract. 2017;26(1):41–49. doi: 10.1159/000450645. Epub 2016 Sep 7. PMID: 27607437; PMCID: PMC5588308

6) Schardt, C., Adams, M. B., Owens, T., Keitz, S., & Fontelo, P. (2007). Utilization of the PICO framework to improve PubMed search for clinical questions. BMC Medical Informatics and Decision Making, 7, 16. doi http://dx.doi.org/10.1186/1472-6947-7

7) van Tulder M, Furlan A, Bombardier C, Bouter L; Editorial Board of the Cochrane Collaboration Back Review Group. Updated method guidelines for systematic reviews in the cochrane collaboration back review group. Spine (Phila Pa 1976). 2003 Jun 15;28(12):1290–9. doi: 10.1097/01.BRS.0000065484.95996.AF. PMID: 12811274.

8) Centre for Reviews and Dissemination. (2009). Systematic reviews: CRD’s guidance for undertaking reviews in health care. University of York. https://www.york.ac.uk/crd/guidance/

9) Furlan AD, Pennick V, Bombardier C, van Tulder M. 2009 updated method guideline for systematic reviews in the Cochrane Back Review Group. Spine. 2009;34(18):1929–41. doi:10.1097/BRS.0b013e3181b1c99f

10) Husereau, D., Drummond, M., Augustovski, F., de Bekker-Grob, E., Briggs, A. H., Carswell, C… & Loder, E. (2022). Consolidated Health Economic Evaluation Reporting Standards 2022 (CHEERS 2022) Explanation and Elaboration: A Report of the ISPOR CHEERS II Good Practices Task Force. Value in Health, 25(1), 10–31. 10.1016/j.jval.2021.10.008

11) Andria Sirur, Sushmitha Jattan, Ardra C Jayan, Aditya Tanwar, Aneesha Sathish, Girish Thunga, Muhammed Rashid, Pooja Gopal Poojari. Cost effectiveness and Cost utility of Complementary and Integrative medicine compared to Alternative, Traditional or Conventional medicine alone in Low- and Middle-Income Countries (LMIC): A Systematic Review Protocol. PROSPERO 2024 Available from https://www.crd.york.ac.uk/PROSPERO/view/CRD42024526580

12) Moher D, Shamseer L, Clarke M, Ghersi D, Liberati A, Petticrew M, Shekelle P, Stewart LA; PRISMA-P Group. Preferred reporting items for systematic review and meta-analysis protocols (PRISMA-P) 2015 statement. Syst Rev. 2015 Jan 1;4(1):1. doi: 10.1186/2046-4053-4-1. PMID: 25554246; PMCID: PMC4320440.

